# Elevated liver enzymes and bilirubin following SARS-CoV-2 infection in children under 10

**DOI:** 10.1101/2022.05.10.22274866

**Authors:** Ellen K. Kendall, Veronica R. Olaker, David C. Kaelber, Rong Xu, Pamela B. Davis

**Affiliations:** Center for Artificial Intelligence in Drug Discovery, Case Western Reserve University School of Medicine, Cleveland, Ohio; The Center for Clinical Informatics Research and Education, The MetroHealth System, Cleveland, Ohio; Center for Community Health Integration, Case Western Reserve University School of Medicine, Cleveland, Ohio

## Abstract

Recently, the Centers for Diseases and Control released a nationwide health alert about an increase in hepatitis cases of unknown origin in children, raising concern about potential sequelae of COVID-19 infection. In this study, we test whether there was increased risk of elevated serum liver enzymes and bilirubin following COVID-19 infection in children. We performed a retrospective cohort study on a nation-wide database of patient electronic health records (EHRs) in the US. The study population comprise 796,369 children between the ages of 1-10 years including 245,675 who had contracted COVID-19 during March 11, 2020 - March 11, 2022 and 550,694 who contracted non-COVID other respiratory infection (ORI) during the same timeframe.

Compared to children infected with other respiratory infections, children infected with COVID-19 infection were at significantly increased risk for elevated AST or ALT (hazard ratio or HR: 2.52, 95% confidence interval or CI: 2.03-3.12) and total bilirubin (HR: 3.35, 95% CI: 2.16-5.18). These results suggest acute and long-term hepatic sequelae of COVID-19 in pediatric patients. Further investigation is needed to clarify if post-COVID-19 related hepatic injury described in this study is related to the current increase in pediatric hepatitis cases of unknown origin.

## Introduction

COVID-19 infection has been associated with hepatic involvement in adults, often manifesting as elevated serum alanine aminotransferase (ALT) and aspartate aminotransferase (AST) levels during infection, as well as direct RNA viral detection in liver samples. ^1,2^ Early evidence from a study in India described 37 pediatric patients presenting with clinical signs of hepatitis after having COVID-19, but how COVID-19 affects the liver in pediatric patients remains unknown. ^3^ Recently, the Centers for Diseases and Control released a nationwide health alert about an increase in hepatitis cases of unknown origin in children, raising concern about potential sequelae of COVID-19 infection. ^4^ In this study, we test whether there was increased risk of elevated serum liver enzymes and bilirubin following COVID-19 infection in children.

## Methods

The TriNetX Analytics Platform was used for data analysis. TriNetX global health collaborative database is a large de-identified database representing over 75 healthcare organizations and 93 million patients across 14 countries and all 50 U.S. states. Studies using TriNetX in this way have been determined to be IRB exempt by the MetroHealth System IRB (Cleveland Ohio).

The study population included 796,369 children between the ages of 1-10 years and was divided into two cohorts. COVID-19 cohort (n = 245,675): with new COVID-19 infection recorded from March 11, 2020 - March 11, 2022. The control cohort (550,694): with non-COVID other respiratory infection (ORI) during the same timeframe. Exclusion criteria included patients with diagnoses of cancer, viral hepatitis, alpha-1-antitrypsin deficiency, and other diseases of the liver. Cohorts were propensity-score matched (1:1 using a nearest neighbor greedy matching) for age, race, ethnicity, pediatric BMI, overweight, and obesity (Table).

Risks of elevated AST (≥ 110) or ALT (≥ 100) and serum total bilirubin (≥ 2) not previously documented in these patients were compared between the COVID-19 and ORI cohorts using hazard ratios (HR) and 95% confidence intervals (CI). Outcomes were reported at 1 month, 3 months, and 6 months following infection. All statistical analyses were conducted in TriNetX Analytics Platform. Details of cohort definitions and statistical analyses were described in eMethods.

## Results

After propensity-score matching, there were 245,161 patients in the COVID-19 cohort and 245,161 in the ORI control cohort (**Table 1**). From 1 day to 6 months after the index event, 269 patients in the COVID-19 cohort and 121 in the ORI cohort had AST ≥ 110 U/L or ALT ≥ 100 U/L (HR: 2.52, 95% CI: 2.03-3.12). 79 patients in the COVID-19 cohort and 27 in the ORI cohort had serum total bilirubin ≥ 2 mg/dL (HR: 3.35, 95% CI: 2.16-5.18) (**Figure 1**). Similar results were obtained comparing 1–4-year-old patients with COVID-19 infection vs ORI at 1 month, 3 months, and 6 months. The specific 1–4-year-old age group was selected, as this is an unvaccinated population and the peak age for hepatitis—as described in the CDC alert.

**Table 1.** Characteristics of the study population before and after propensity-score matching

| Characteristics | Before Matching |  |  | After Matching |  |  |
| --- | --- | --- | --- | --- | --- | --- |
|  | Cohort, No. (%) |  |  | Cohort, No. (%) |  |  |
|  | COVID-19 | ORI | SMD | COVID-19 | ORI | SMD |
| Total number | 245,675 | 550,694 |  | 245,161 | 245,161 |  |
| Current age, mean (SD), y | 6.0 (+/- 3.1) | 5.3 (+/- 3.0) | 0.222 | 6.0 (+/- 3.1) | 6.0 (+/- 3.1) | 0.002 |
| Age at index, mean (SD), y | 5.1 (+/- 3.0) | 4.4 (+/- 2.9) | 0.243 | 5.1 (+/- 3.0) | 5.1 (+/- 3.0) | 0.001 |
| Sex - female | 114,954(46.8) | 261,837(47.5) | 0.015 | 114,773(46.8%) | 115,203(47.0) | 0.004 |
| Sex - male | 130,672(53.2) | 288,782(52.4) | 0.015 | 130,341(53.2) | 129,923(53.0) | 0.003 |
| Ethnicity - Hispanic/Latinx | 59,525(24.2) | 90,193(16.4) | 0.196 | 59,061(24.1) | 59,556(24.3) | 0.005 |
| Ethnicity - Not Hispanic/Latinx | 140,744(57.3) | 294,765(53.5) | 0.076 | 140,694(57.4) | 140,181(57.2) | 0.004 |
| Ethnicity - Unknown | 45,406(18.5) | 165,736(30.1) | 0.273 | 45,406(18.5) | 45,424(18.5) | <0.001 |
| Race - American Indian or Alaska Native | 855(0.3) | 2,295(0.4) | 0.011 | 855(0.3) | 795(0.3) | 0.004 |
| Race - Asian | 6,808(2.8) | 14,717(2.7) | 0.006 | 6,800(2.8) | 6,764(2.8) | 0.001 |
| Race - Black or African American | 44,963(18.3) | 87,405(15.9) | 0.065 | 44,897(18.3) | 44,509(18.2) | 0.004 |
| Race - Native Hawaiian or Other Pacific Islander | 281(0.1) | 798(0.1) | 0.008 | 281(0.1) | 242(0.1) | 0.005 |
| Race - Unknown | 62,150(25.3) | 177,877(32.3) | 0.155 | 62,106(25.3) | 62,655(25.6) | 0.005 |
| Race - White | 130,618(53.2) | 267,602(48.6) | 0.092 | 130,222(53.1) | 130,196(53.1) | <0.001 |
| Overweight and Obesity | 9,724(4.0) | 16,103(2.9) | 0.057 | 9,594(3.9) | 9,640(3.9) | 0.001 |
| Body mass index [BMI] Pediatric | 22,985(9.4) | 70,988(12.9) | 0.113 | 22,985(9.4) | 22,998(9.4) | <0.001 |

**Figure 1.**
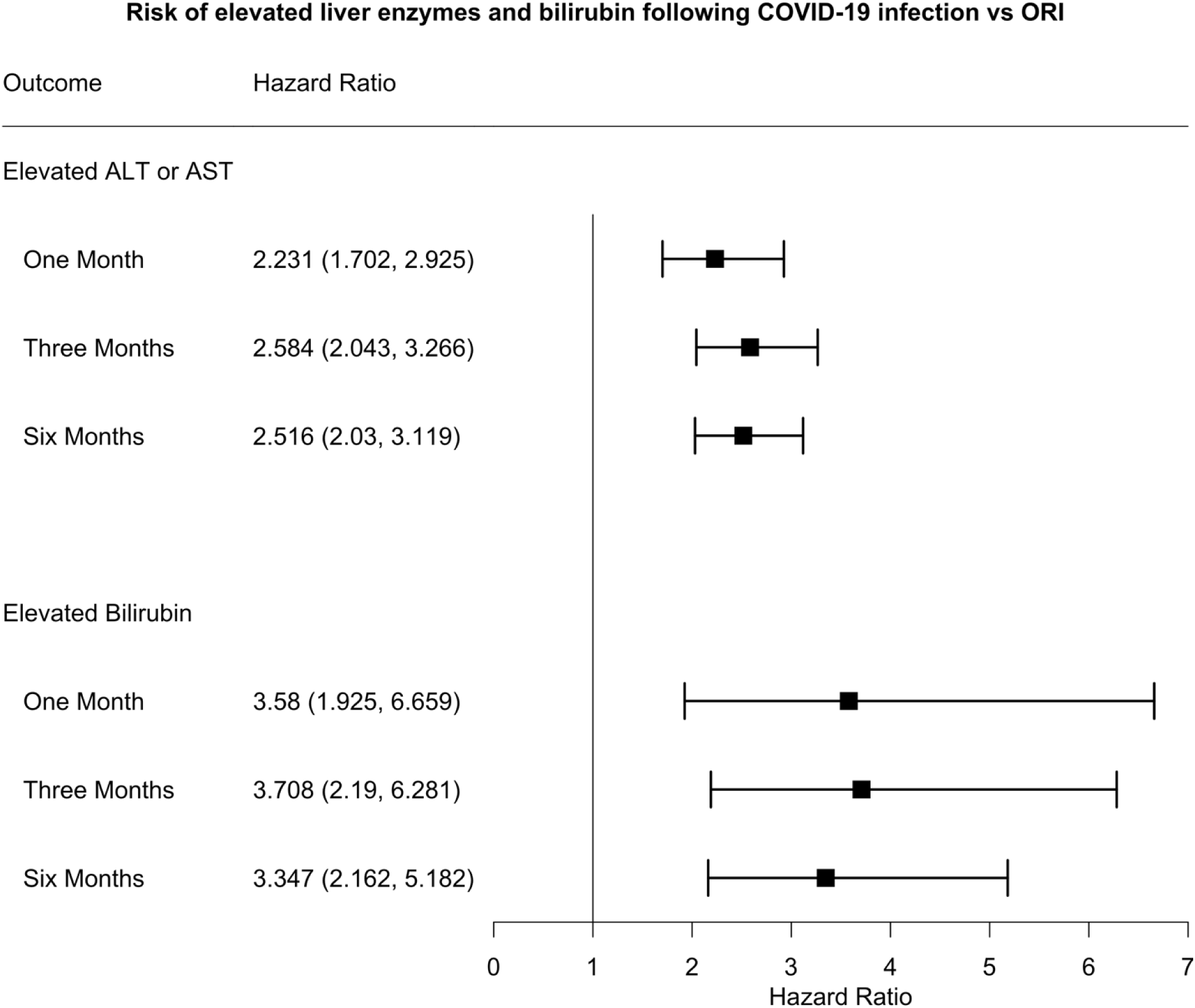
Comparison of risks for elevated liver enzymes and bilirubin within 1, 3, 6 months following COVID-19 or non-COVID other respiratory infection (ORI) between propensity-score matching cohorts.

## Discussion

Following COVID-19 infection, there was a higher risk of elevated AST or ALT and total bilirubin at 1, 3, and 6 months in comparison to children infected with other respiratory infections. These results suggest acute and long-term hepatic sequelae of COVID-19 in pediatric patients, raising concern for long term clinical outcomes following a COVID-19 related hepatic injury. Study limitations include potential biases due to the observational nature of this study, and the need to confirm these results in other datasets and populations. Further investigation is needed to clarify if post-COVID-19 related hepatic injury described in this study is related to the current increase in pediatric hepatitis cases of unknown origin.

## Supporting information

eMethod

## Data Availability

All data produced in the present work are contained in the manuscript

*Concept and design: PBD, RX, EKK, VRO*

*Acquisition, analysis, or interpretation of data:* EKK, VRO, PBD, RX

*Drafting of the manuscript: EKK, VRO*

*Critical revision of the manuscript for important intellectual content:* RX, PBD

*Statistical analysis:* EKK, VO

*Obtained funding:* PBD, RX

*Administrative, technical, or material support:* PBD, RX, DCK

*Supervision:* PBD, DCK, RX

