## Supplementary material for "Elevated liver enzymes and bilirubin following SARS-CoV-2 infection in children under 10": eMethod

**eMethods.**

**TriNetX Database and Statistical Analysis Description of TriNetX database**: The data used in this study was accessed on April 28^th^, 2022 from the TriNetX Global Collaborative Network, which provides access to electronic medical records (diagnoses, procedures, medications, laboratory values, genomic information) from over 93 million patients from 74 healthcare organizations. Data are de-identified per criteria from the Health Insurance Portability and Accountability Act (HIPAA), Section §164.514(a) of the HIPAA Privacy Rule. The MetroHealth System, Cleveland, Ohio, IRB has determined any research using TriNetX, is not Human Subject Research and therefore exempt from IRB review.

The TriNetX platform de-identifies and aggregates electronic health record (EHR) data from 74 contributing healthcare systems, most of which are large academic medical institutions with both inpatient and outpatient facilities at multiple locations, across 50 states in the US and 14 countries. Patient EHR data includes information from hospitals, primary care, and specialty treatment providers, covering diverse geographic locations, age groups, racial and ethnic groups, income levels and insurance types including commercial insurances, governmental insurance (Medicare and Medicaid), self-pay/uninsured, worker compensation insurance, and military/VA insurance, among others. Race and ethnicity data in TriNetX is derived from self-reports, which are then mapped to the following categories: (1) Race: Asian, American Indian or Alaskan Native, Black or African American, Native Hawaiian or Other Pacific Islander, White, Unknown race; and (2) Ethnicity: Hispanic or Latino, Not Hispanic or Latino, Unknown Ethnicity.

**Statistical analysis:** The status of SARS-CoV-2 infection was based on the lab-test confirmed presence or the International Classification of Diseases (ICD-10) diagnosis codes (complete cohort terms defined below). The outcome measure of elevated AST or ALT was determined by the presence of serum or plasma AST ≥ 110 U/L or serum or plasma ALT ≥ 100 U/L as the most recent lab value. The outcome measure of elevated bilirubin was determined by the presence of serum, plasma, or blood total bilirubin ≥ 2 mg/dL as the most recent lab value. SARS-CoV-2 infections that were diagnosed between March 11, 2020 and March 11^th^, 2022 were recorded for children aged 1-10 years. We additionally ran an analysis for children aged 1-4 years. We established a cohort of children diagnosed with other respiratory infection (cohort terms below) who were never diagnosed with SARS-CoV2 during the same time period. Each cohort was matched with the cohort of children with SARS-CoV2 infection by the TriNetX built‐in propensity score matching function (1:1 matching using a nearest neighbor greedy matching algorithm with a caliper of 0.25 times the standard deviation).  Risk of new elevated AST/ALT or bilirubin following date of infection was then compared for the SARS-CoV2 cohort to the other respiratory infection cohort (or routine visit cohort) using hazard ratios and 95% confidence intervals. Kaplan-Meier analysis was used to estimate the probability of clinical outcomes. Cox’s proportional hazards model was used to compare the two matched cohorts. The proportional hazard assumption was tested using the generalized Schoenfeld approach. For sensitivity analysis, we also created one additional control cohort of children who had routine encounters with the health care system but were never diagnosed with SARS-CoV2. Risks for new elevated AST/ALT and bilirubin were compared between children with SARS-CoV-2 infection and these control cohorts as described above. The TriNetX Platform calculates the hazard ratios and associated confidence intervals, using R's Survival package v3.2-3. For generating hazard ratios, TriNetX sets robust=FALSE using the R survival package, but it does not take into account potential clustering of COVID-19 cases within the healthcare organizations or specific geolocations, a potential weakness or confounding factor in the analysis.

The full ICD-10 code list for defining the COVID-19 cohort is listed below. This list was adapted from reference 6 and includes a mix of diagnostic and laboratory codes. This same list was used as exclusion criteria for the other respiratory infection cohort.

| UMLS:ICD10CM:U07.2 | COVID-19, virus not identified (WHO) (between 1 and 10 years old at event) |
| --- | --- |
| UMLS:ICD10CM:U07.1 | COVID-19 (between 1 and 10 years old at event) |
| UMLS:ICD10CM:J12.81 | Pneumonia due to SARS-associated coronavirus (between 1 and 10 years old at event) |
| UMLS:ICD10CM:B97.29 | Other coronavirus as the cause of diseases classified elsewhere (between 1 and 10 years old at event) |
| UMLS:ICD10CM:B34.2 | Coronavirus infection, unspecified (between 1 and 10 years old at event) |
| TNX:9088 | SARS coronavirus 2 and related RNA [Presence] (between 1 and 10 years old at event; labResult: Positive) |
| UMLS:LNC:94500-6 | SARS-CoV-2 (COVID-19) RNA [Presence] in Respiratory specimen by NAA with probe detection (between 1 and 10 years old at event; labResult: Positive) |
| UMLS:LNC:94309-2 | SARS-CoV-2 (COVID-19) RNA [Presence] in Specimen by NAA with probe detection (between 1 and 10 years old at event; labResult: Positive) |
| UMLS:LNC:94534-5 | SARS-CoV-2 (COVID-19) RdRp gene [Presence] in Respiratory specimen by NAA with probe detection (between 1 and 10 years old at event; labResult: Positive) |
| UMLS:LNC:94565-9 | SARS-CoV-2 (COVID-19) RNA [Presence] in Nasopharynx by NAA with non-probe detection (between 1 and 10 years old at event; labResult: Positive) |
| UMLS:LNC:94315-9 | SARS-related coronavirus E gene [Presence] in Specimen by NAA with probe detection (between 1 and 10 years old at event; labResult: Positive) |
| UMLS:LNC:94759-8 | SARS-CoV-2 (COVID-19) RNA [Presence] in Nasopharynx by NAA with probe detection (between 1 and 10 years old at event; labResult: Positive) |
| UMLS:LNC:94316-7 | SARS-CoV-2 (COVID-19) N gene [Presence] in Specimen by NAA with probe detection (between 1 and 10 years old at event; labResult: Positive) |
| UMLS:LNC:94559-2 | SARS-CoV-2 (COVID-19) ORF1ab region [Presence] in Respiratory specimen by NAA with probe detection (between 1 and 10 years old at event; labResult: Positive) |
| UMLS:LNC:95209-3 | SARS-CoV+SARS-CoV-2 (COVID-19) Ag [Presence] in Respiratory specimen by Rapid immunoassay (between 1 and 10 years old at event; labResult: Positive) |
| UMLS:LNC:94558-4 | SARS-CoV-2 (COVID-19) Ag [Presence] in Respiratory specimen by Rapid immunoassay (between 1 and 10 years old at event; labResult: Positive) |
| UMLS:LNC:95608-6 | SARS-CoV-2 (COVID-19) RNA [Presence] in Respiratory specimen by NAA with non-probe detection (between 1 and 10 years old at event; labResult: Positive) |
| UMLS:LNC:96119-3 | SARS-CoV-2 (COVID-19) Ag [Presence] in Upper respiratory specimen by Immunoassay (between 1 and 10 years old at event; labResult: Positive) |
| UMLS:LNC:94763-0 | SARS-CoV-2 (COVID-19) [Presence] in Specimen by Organism specific culture (between 1 and 10 years old at event; labResult: Positive) |
| UMLS:LNC:94533-7 | SARS-CoV-2 (COVID-19) N gene [Presence] in Respiratory specimen by NAA with probe detection (between 1 and 10 years old at event; labResult: Positive) |
| UMLS:LNC:94760-6 | SARS-CoV-2 (COVID-19) N gene [Presence] in Nasopharynx by NAA with probe detection (between 1 and 10 years old at event; labResult: Positive) |
| UMLS:LNC:95406-5 | SARS-CoV-2 (COVID-19) RNA [Presence] in Nose by NAA with probe detection (between 1 and 10 years old at event; labResult: Positive) |
| UMLS:LNC:94845-5 | SARS-CoV-2 (COVID-19) RNA [Presence] in Saliva (oral fluid) by NAA with probe detection (between 1 and 10 years old at event; labResult: Positive) |
| UMLS:LNC:94758-0 | SARS-related coronavirus E gene [Presence] in Respiratory specimen by NAA with probe detection (between 1 and 10 years old at event; labResult: Positive) |
| UMLS:LNC:96763-8 | SARS-CoV-2 (COVID-19) E gene [Presence] in Respiratory specimen by NAA with probe detection (between 1 and 10 years old at event; labResult: Positive) |
| UMLS:LNC:94502-2 | SARS-related coronavirus RNA [Presence] in Respiratory specimen by NAA with probe detection (between 1 and 10 years old at event; labResult: Positive) |
| UMLS:LNC:94314-2 | SARS-CoV-2 (COVID-19) RdRp gene [Presence] in Specimen by NAA with probe detection (between 1 and 10 years old at event; labResult: Positive) |
| UMLS:LNC:96123-5 | SARS-CoV-2 (COVID-19) RdRp gene [Presence] in Upper respiratory specimen by NAA with probe detection (between 1 and 10 years old at event; labResult: Positive) |
| UMLS:LNC:97097-0 | SARS-CoV-2 (COVID-19) Ag [Presence] in Upper respiratory specimen by Rapid immunoassay (between 1 and 10 years old at event; labResult: Positive) |
| UMLS:LNC:94647-5 | SARS-related coronavirus RNA [Presence] in Specimen by NAA with probe detection (between 1 and 10 years old at event; labResult: Positive) |
| UMLS:LNC:95409-9 | SARS-CoV-2 (COVID-19) N gene [Presence] in Nose by NAA with probe detection (between 1 and 10 years old at event; labResult: Positive) |

The list of ICD-10 diagnostic codes used as inclusion criteria to define the other respiratory infection cohort are below.

| UMLS:ICD10CM:J00 | Acute nasopharyngitis [common cold] (between 1 and 10 years old at event) |
| --- | --- |
| UMLS:ICD10CM:J04 | Acute laryngitis and tracheitis (between 1 and 10 years old at event) |
| UMLS:ICD10CM:J05 | Acute obstructive laryngitis [croup] and epiglottitis (between 1 and 10 years old at event) |
| UMLS:ICD10CM:J03 | Acute tonsillitis (between 1 and 10 years old at event) |
| UMLS:ICD10CM:J02 | Acute pharyngitis (between 1 and 10 years old at event) |
| UMLS:ICD10CM:J09-J18 | Influenza and pneumonia (between 1 and 10 years old at event) |
| UMLS:ICD10CM:J06 | Acute upper respiratory infections of multiple and unspecified sites (between 1 and 10 years old at event) |
| UMLS:ICD10CM:J01 | Acute sinusitis (between 1 and 10 years old at event) |
| UMLS:ICD10CM:J20-J22 | Other acute lower respiratory infections (between 1 and 10 years old at event) |

Exclusion criteria for both cohorts included liver disease, alpha-1 antitrypsin deficiency, cancer, and viral hepatitis, and these were excluded for being present at any point in time. The ICD-10 code list is below.

UMLS:ICD10CM:E88.01	Alpha-1-antitrypsin deficiency

UMLS:ICD10CM:B15-B19	Viral hepatitis

UMLS:ICD10CM:K76	Other diseases of liver

UMLS:ICD10CM:K75.8	Other specified inflammatory liver diseases

UMLS:ICD10CM:K75.4	Autoimmune hepatitis

UMLS:ICD10CM:K75.3	Granulomatous hepatitis, not elsewhere classified

UMLS:ICD10CM:K75.2	Nonspecific reactive hepatitis

UMLS:ICD10CM:K75.1	Phlebitis of portal vein

UMLS:ICD10CM:K75.0	Abscess of liver

UMLS:ICD10CM:K74	Fibrosis and cirrhosis of liver

UMLS:ICD10CM:K73	Chronic hepatitis, not elsewhere classified

UMLS:ICD10CM:K72	Hepatic failure, not elsewhere classified

UMLS:ICD10CM:K71	Toxic liver disease

UMLS:ICD10CM:K70	Alcoholic liver disease

UMLS:ICD10CM:C00-D49	Neoplasms

One final exclusion criterion used in both analyses was a diagnostic code within code K75: Other inflammatory liver disease. Code K75.9 Inflammatory liver disease, unspecified was excluded up until March 10, 2020, which is the date before the index event time window began. This code was allowed to occur within the index time window and analysis follow-up period in case it may have been utilized as a clinical identification for children with post-viral elevated liver enzymes.
